# Epidemiology, clinical course, and outcomes of critically ill adults with COVID-19 in New York City: a prospective cohort study

**DOI:** 10.1101/2020.04.15.20067157

**Authors:** Matthew J. Cummings, Matthew R. Baldwin, Darryl Abrams, Samuel D. Jacobson, Benjamin J. Meyer, Elizabeth M. Balough, Justin G. Aaron, Jan Claassen, LeRoy E. Rabbani, Jonathan Hastie, Beth R. Hochman, John Salazar-Schicchi, Natalie H. Yip, Daniel Brodie, Max R. O’Donnell

**Author notes:** **Corresponding Author:** Max R. O’Donnell, MD, MPH, Division of Pulmonary, Allergy, and Critical Care Medicine, Columbia University Medical Center, 622 West 168^th^ St, PH 8E-101, New York, NY, 10032, USA.

## Abstract

**Background:** Nearly 30,000 patients with coronavirus disease-2019 (COVID-19) have been hospitalized in New York City as of April 14^th^, 2020. Data on the epidemiology, clinical course, and outcomes of critically ill patients with COVID-19 in this setting are needed.

**Methods:** We prospectively collected clinical, biomarker, and treatment data on critically ill adults with laboratory-confirmed-COVID-19 admitted to two hospitals in northern Manhattan between March 2^nd^ and April 1^st^, 2020. The primary outcome was in-hospital mortality.

Secondary outcomes included frequency and duration of invasive mechanical ventilation, frequency of vasopressor use and renal-replacement-therapy, and time to clinical deterioration following hospital admission. The relationship between clinical risk factors, biomarkers, and in-hospital mortality was modeled using Cox-proportional-hazards regression. Each patient had at least 14 days of observation.

**Results:** Of 1,150 adults hospitalized with COVID-19 during the study period, 257 (22%) were critically ill. The median age was 62 years (interquartile range [IQR] 51-72); 170 (66%) were male. Two-hundred twelve (82%) had at least one chronic illness, the most common of which were hypertension (63%; 162/257) and diabetes mellitus (36%; 92/257). One-hundred-thirty-eight patients (54%) were obese, and 13 (5%) were healthcare workers. As of April 14^th^, 2020, in-hospital mortality was 33% (86/257); 47% (122/257) of patients remained hospitalized. Two-hundred-one (79%) patients received invasive mechanical ventilation (median 13 days [IQR 9-17]), and 54% (138/257) and 29% (75/257) required vasopressors and renal-replacement-therapy, respectively. The median time to clinical deterioration following hospital admission was 3 days (IQR 1-6). Older age, hypertension, chronic lung disease, and higher concentrations of interleukin-6 and d-dimer at admission were independently associated with in-hospital mortality.

**Conclusions:** Critical illness among patients hospitalized with COVID-19 in New York City is common and associated with a high frequency of invasive mechanical ventilation, extra-pulmonary organ dysfunction, and substantial in-hospital mortality.

## Introduction

As of April 14^th^, 2020, nearly 580,000 cases of coronavirus disease-2019 (COVID-19) associated with SARS-CoV-2 infection had been reported in the United States.^1^ Of these, over 202,000 were reported in New York State.^2^ In New York City, over 110,000 cases were reported, of which approximately 30,000 (28%) had been hospitalized.^2^

Available data suggest that 5-20% of patients with COVID-19 develop critical illness that is characterized primarily by the acute respiratory distress syndrome (ARDS).^3-6^ Although the clinical spectrum of severe COVID-19 has been characterized in reports from China and Italy,^3-8^ understanding of COVID-19-related critical illness in the U.S. has been limited to small case-series from Washington state.^9,10^ Here, we characterize the epidemiology, clinical course, and risk factors for in-hospital mortality among a large cohort of adults with COVID-19-related critical illness admitted to two hospitals in New York City during the first 30 days of the city’s outbreak.

## Methods

### Study Setting, Design, and Participants

This prospective observational cohort study was conducted at two New York-Presbyterian hospitals affiliated with Columbia University Irving Medical Center in northern Manhattan. The two hospitals, a 700-bed tertiary referral hospital, and a 230-bed community-based hospital, included 117 and 12 intensive care unit (ICU) beds, respectively, prior to the COVID-19 pandemic. We prospectively identified adult patients (age ≥18 years) admitted to both hospitals from March 2^nd^ to April 1^st^, 2020, who were diagnosed with laboratory-confirmed COVID-19 and who were critically ill with acute hypoxemic respiratory failure, defined as those receiving mechanical ventilation (invasive or non-invasive) or high-level supplemental oxygen via high-flow nasal cannula or non-rebreathing face mask at a flow rate of 15 liters/minute or greater at or during hospitalization. Laboratory confirmation of SARS-CoV-2 infection was performed using real-time reverse-transcription polymerase-chain-reaction (rtRT-PCR) testing of naso- and/or oro-pharyngeal swab samples by the New York City Department of Health from March 2^nd^-March 10^th^, 2020, after which testing was performed using rtRT-PCR in the clinical microbiological laboratory of the referral hospital. We identified critically ill patients with COVID-19 through daily review of hospital admission logs in the electronic medical record.

### Data Collection

We reviewed electronic medical records, laboratory results, and radiographic findings for all admitted patients with critical illness and laboratory-confirmed COVID-19. Using a standardized case record form developed by the International Severe Acute Respiratory and Emerging Infection Consortium and World Health Organization (WHO),^11^ we recorded data on demographics, known medical history and co-morbidities, illness onset and symptoms, vital signs and biochemical studies performed within 24 hours of diagnosis of acute respiratory failure. We also recorded concentrations of plasma- and serum-based biomarkers drawn at or within 48 hours of hospital admission, including C-reactive protein, d-dimer, ferritin, high-sensitivity troponin, procalcitonin, and interleukin (IL)-6. We prospectively collected data on management interventions delivered during hospitalization including initiation and duration of mechanical ventilation, administration of advanced therapies for acute respiratory failure (neuromuscular blocking agents, inhaled pulmonary vasodilators, prone-positioning, and extracorporeal membrane oxygenation), vasopressor agents, renal replacement therapy, anti-bacterial, anti-viral, and immunomodulatory agents (interleukin-6-receptor antagonists and corticosteroids).

### Outcomes

The primary outcome was the rate of in-hospital death. Follow-up time was right-censored on April 14^th^, 2020. Secondary outcomes included frequency and duration of invasive mechanical ventilation, frequency of vasopressor use and renal replacement therapy, and the time to in-hospital clinical deterioration following admission, defined as an increase of one-point from baseline on a 7-point ordinal scale. This scale, designed to assess clinical status over time, was based on that recommended by WHO for use in clinical research among hospitalized patients with COVID-19 (Table S1 in data supplement).^12^

### Statistical analyses

Continuous variables were expressed as means (standard deviation) and medians (interquartile ranges). Categorical variables were summarized as counts and percentages. Missing data was rare and not imputed. We created Kaplan-Meier survival plots and used the log rank test to compare survival patterns by co-morbidity. We estimated hazard ratios for death using Cox proportional-hazards models. We measured time-to-event in days from the date of hospital admission to the date of in-hospital death or hospital discharge alive. Follow-up time was right-censored on April 14^th^, 2020. We included age, sex, duration of symptoms prior to hospital presentation, severe obesity (defined as body-mass-index ≥35), and co-morbidities (hypertension, chronic cardiovascular, pulmonary, and kidney disease and diabetes mellitus) as independent variables in our Cox models. We also included serum IL-6 and plasma d-dimer concentrations as independent variables in our models because there is emerging evidence of dysregulated immune activation and coagulopathy in patients with severe COVID-19, and interest in treating this patient population with targeted immunomodulatory therapies and anti-coagulation.^13,14^ We confirmed the proportional hazards assumption of the Cox models using the Schoenfeld residuals test. All analyses were performed using Stata (version 16, StataCorp, College Station, TX, USA).

### Ethics Statement

This study was approved by the institutional review board at Columbia University Irving Medical Center. The requirement for written informed consent was waived because of the study design and ongoing public health emergency.

## Results

### Patient Characteristics

Between March 2^nd^ and April 1^st^, 2020, 1,150 adults were admitted to both hospitals with laboratory-confirmed COVID-19, of which 257 (22%) were critically ill (Table 1). The median period of observation following hospital admission was 15 days (IQR 9-19). The median age was 62 years (interquartile range [IQR] 51-72); 170 patients (66%) were male, 158 (62%) were Hispanic or Latino, and 13 (5%) were healthcare workers (HCWs). Two-hundred-twelve (82%) had at least one chronic illness, of which hypertension and diabetes mellitus were the most common. The mean BMI was 30.8 (±7.7); 68 (26%) of patients had a BMI ≥ 35 (Table 1). Patients presented a mean of 6 (±5) days after symptom onset. The most common presenting symptoms were shortness of breath, fever, cough, myalgia, and diarrhea. Nearly all (98%; 252/257) patients had infiltrates present on initial chest radiograph (Table 1). The median lactate concentration was 1.5 (IQR 1.1-2.2). Median serum creatinine was 1.5 (IQR 1.9-2.4) and 87% (189/218) of patients with urinalysis performed had proteinuria. Lymphocytopenia was common as was mild elevation of aspartate aminotransferase. Concentrations of CRP (available in 98% of patients; 253/257), ferritin (98%; 253/257), d-dimer (95%; 244/257), high-sensitivity troponin (99%; 254/257), procalcitonin (99%; 255/257), and IL-6 (92%; 237/257) were elevated in most patients (Table 2).

**Table 1:** Patient Characteristics.

| Patient characteristic | Study population, n=257 |
| --- | --- |
| Male sex, n (%) | 170/257 (66%) |
| Age group, (years), n (%) |  |
| 20-29 | 8/257 (3%) |
| 30-39 | 19/257 (8%) |
| 40-49 | 28/257 (11%) |
| 50-59 | 52/257 (20%) |
| 60-69 | 69/257 (27%) |
| 70-79 | 52/257 (20%) |
| 80-89 | 23/257 (9%) |
| ≥90 | 6/257 (2%) |
| Age, years, median (IQR) | 62 (51-72) |
| Race or ethnic group, n (%) |  |
| Hispanic or Latino | 158/257 (62%) |
| Black or African American | 49/257 (19%) |
| White | 32/257 (12%) |
| Asian | 9/257 (4%) |
| Other | 5/257 (3%) |
| Body mass index, kg/m<sup>2</sup>, mean (±SD) | 30.8 (±7.7) |
| Body mass index ≥ 30, n (%) | 119/257 (46%) |
| Body mass index ≥ 35, n (%) | 68/257 (26%) |
| Body mass index ≥ 40, n (%) | 33/257 (13%) |
| Employed as healthcare worker, n (%) | 13/257 (5%) |
| Co-existing disorder, n (%) |  |
| Hypertension | 162/257 (63%) |
| Diabetes mellitus | 92/257 (36%) |
| Chronic cardiovascular disease excluding hypertension<sup>a</sup> | 49/257 (19%) |
| Chronic kidney disease<sup>b</sup> | 37/257 (14%) |
| Current or former smoker | 33/257 (13%) |
| Chronic obstructive pulmonary or interstitial lung disease | 24/257 (9%) |
| Chronic neurological disease<sup>c</sup> or dementia | 24/257 (9%) |
| Asthma | 21/257 (8%) |
| Active solid or hematologic malignancy or dysplasia | 18/257 (7%) |
| Solid organ transplant recipient | 10/257 (4%) |
| Human immunodeficiency virus infection | 8/257 (3%) |
| Liver cirrhosis<sup>d</sup> | 5/257 (2%) |
| Duration of illness prior to presentation, days, mean (±SD) | 6 (±5) |
| Symptoms reported, n (%) |  |
| Shortness of breath | 190/257 (74%) |
| Fever | 183/257 (71%) |
| Cough | 169/257 (66%) |
| Myalgia | 67/257 (26%) |
| Diarrhea | 32/257 (12%) |
| Rhinorrhea | 19/257 (7%) |
| Sore throat | 15/257 (6%) |
| Headache | 10/257 (4%) |
| Vital signs at hospital presentation |  |
| Temperature, celsius, mean (±SD) | 38.3 (± 6.0) |
| Heart rate, beats/minute, mean (±SD) | 101 (± 20) |
| Respiratory rate, breaths/minute, mean (±SD) | 22 (± 6) |
| Systolic blood pressure, mmHg, mean (±SD) | 129 (± 23) |
| Oxygen saturation, %, mean (±SD) | 89 (± 10) |
| Altered mental status, n (%) | 23 (9%) |
| Infiltrates present on initial chest radiograph, n (%) | 252/257 (98%) |

Abbreviations: IQR: interquartile range, SD: standard deviation
Legend: <sup>a</sup>Coronary artery disease or congestive heart failure; <sup>b</sup>Chronic kidney disease of any stage; <sup>c</sup>Chronic neurodegenerative disease or history of stroke; <sup>d</sup>Liver cirrhosis of any Child-Pugh class.

**Table 2:** Biochemical and Biomarker Values.

| Variable | Study population, n=257 |
| --- | --- |
| Lactate, mmol/L, median (IQR) | 1.5 (1.1-2.2) |
| Creatinine, mg/dl, median (IQR) | 1.5 (0.9-2.4) |
| Proteinuria, n (%) | 189/218 (87%) |
| Urine protein concentration, mg/dl, median (IQR) | 100 (30-300) |
| White blood cell count, x 10<sup>3</sup>/μL, median (IQR) | 9.8 (6.6-12.7) |
| Lymphocyte count, x 10<sup>3</sup>/μL, median (IQR) | 0.8 (0.6-1.2) |
| Platelet count, x 10<sup>3</sup>/μL, median (IQR) | 199 (148-270) |
| Bilirubin, mg/dl, median (IQR)<sup>a</sup> | 0.6 (0.4-0.8) |
| AST, U/L, median (IQR)<sup>a</sup> | 61 (42-104) |
| ALT, U/L, median (IQR)<sup>a</sup> | 39 (27-67) |
| Creatine kinase, U/L, median (IQR)<sup>b</sup> | 236 (103-646) |
| Prothrombin time, U/L, median (IQR) | 14.7 (14-16) |
| Interleukin-6, pg/ml<sup>c</sup> |  |
| Mean (±SD) | 65 (±102) |
| Median (IQR) | 23 (10-55) |
| Range | 5-572 |
| High-sensitivity C-reactive protein, mg/l<sup>d</sup> |  |
| Mean (±SD) | 165 (±95) |
| Median (IQR) | 158 (92-254) |
| Range | 4-447 |
| Ferritin, ng/ml<sup>d</sup> |  |
| Mean (±SD) | 1651 (±3229) |
| Median (IQR) | 924 (472-1789) |
| Range | 11-42,406 |
| D-dimer, ng/ml<sup>e</sup> |  |
| Mean (±SD) | 4.4 (±15.7) |
| Median (IQR) | 1.6 (0.9-3.5) |
| Range | 0.3-236 |
| High-sensitivity Troponin-T, ng/L<sup>f</sup> |  |
| Mean (±SD) | 88 (±400) |
| Median (IQR) | 19 (9-52) |
| Range | 2-6,051 |
| Procalcitonin, ng/ml<sup>g</sup> |  |
| Mean (±SD) | 5.5 (±35) |
| Median (IQR) | 0.35 (0.17-1.1) |
| Range | 0.04-400 |

Abbreviations: IQR: interquartile range, SD: standard deviation.
Legend: Data available for <sup>a</sup>256, <sup>b</sup>223, <sup>c</sup>237, <sup>d</sup>253, <sup>e</sup>244, <sup>f</sup>254, <sup>g</sup>255 patients.

### Clinical Management and Outcomes

During hospitalization, 115 (45%), 12 (5%), and 3 (1%) of patients received respiratory support via non-rebreathing oxygen face mask, high-flow nasal cannula, and non-invasive ventilation, respectively (Table 3). Two-hundred-one patients (78%) received invasive mechanical ventilation (IMV) for a median of 13 days (IQR 9-17). Survivors and non-survivors had a median of 14 (12-18) and 5 (IQR 3-10) days of IMV, respectively. Sixty-percent (69/115) of patients who initially received respiratory support via non-rebreathing oxygen face mask, high-flow nasal cannula, or non-invasive ventilation ultimately received IMV. On the first day of critical illness, the median sequential organ failure assessment (SOFA) score^15^ was 11 (IQR 8-13) and the median value of the lowest ratio of partial pressure of arterial oxygen (P_a_O_2_) to the fraction of inspired oxygen (F_i_O_2_) recorded on this day was 129 (IQR 80-203). The median value of the highest positive end-expiratory pressure (PEEP) prescribed during the first 24 hours of IMV was 15 cm H_2_O (IQR 12-18) (Table 3). Among patients receiving IMV, advanced therapies for acute respiratory failure administered during hospitalization included early neuromuscular blockade (25%; 51/201), inhaled nitric oxide (11%, 22/201), prone-positioning ventilation (16%; 32/201), and extracorporeal membrane oxygenation (2%, 5/201). Over half (54%; 138/257) of patients required vasopressor support and nearly a third (29%; 75/257) required renal replacement therapy (RRT). Most patients received anti-viral agents (hydroxychloroquine [72%; 185/257] and remdesivir [9%; 23/257]) and anti-bacterial agents (89%; 229/257). Twenty-six percent (68/257) of patients received corticosteroids and 23% of patients (60/257) received anti-IL-6 receptor antagonists.

**Table 3:** Clinical management and outcomes.

| **Organ Dysfunction and Organ Support and Targeted Therapies Received During Hospitalization** | **Study population**<br>**n=257** |
| --- | --- |
| Sequential Organ Failure Assessment score,<sup>15</sup> median (IQR)<sup>a</sup> | 11 (8-13) |
| Lowest P<sub>a</sub>O<sub>2</sub> to F<sub>i</sub>O<sub>2</sub> ratio on day 1, mmHg, median (IQR)<sup>b,c</sup> | 129 (80-203) |
| **Respiratory support, n (%)** |  |
| Non-rebreathing oxygen face mask | 115/257 (45%) |
| High-flow nasal cannula oxygen therapy | 12/257 (5%) |
| Non-invasive ventilation | 3/257 (1%) |
| Invasive mechanical ventilation (IMV) | 201/257 (79%) |
| Duration, days, median (IQR) | 13 (9-17) |
| Highest PEEP during first 24 hours of IMV, cm H<sub>2</sub>O, median (IQR) | 15 (12-18) |
| Highest F<sub>i</sub>O<sub>2</sub> during first 24 hours of IMV, %, median (IQR) | 100 (80-100) |
| **Advanced therapies for acute respiratory failure, n (%)<sup>d</sup>** |  |
| Early neuromuscular blockade<sup>e</sup> | 51/201 (25%) |
| Inhaled nitric oxide | 22/201 (11%) |
| Prone-positioning ventilation | 32/201 (16%) |
| Extracorporeal membrane oxygenation | 5/201 (2%) |
| **Vasopressors, n (%)** | 138/257 (54%) |
| **Renal replacement therapy, n (%)** | 75/257 (29%) |
| **Anti-viral agent, n (%)** |  |
| Hydroxychloroquine | 185/257 (72%) |
| Remdesivir | 23/257 (9%) |
| **Anti-bacterial agent, n (%)** | 229/257 (89%) |
| **Immunomodulatory agent, n (%)** |  |
| Corticosteroid | 68/257 (26%) |
| Interleukin-6 receptor antagonis | 60/257 (23%) |
| **Outcomes, n (%)** |  |
| Died in-hospital | 86/257 (33%) |
| Duration of hospitalization prior to death, days, median (IQR) | 8 (5-12) |
| Hospitalized | 122/257 (47%) |
| Transferred to another hospital | 4/257 (2%) |
| Discharged alive | 45/257 (18%) |
| New requirement for supplemental oxygen at discharge, n (%) | 9/45 (20%) |

Abbreviations: F<sub>i</sub>O<sub>2</sub>: fraction of inspired oxygen, P<sub>a</sub>O<sub>2</sub>: partial pressure of arterial oxygen, PEEP: positive end-expiratory pressure, IQR: interquartile range, SD: standard deviation.
Legend: <sup>a</sup>Determinable for 221 patients; mental status assessment of alert, responsive to voice, pain, or unresponsive converted to Glasgow Coma Scale for calculation<sup>15</sup>; <sup>b</sup>Determinable for 222 patients; <sup>c</sup>F<sub>i</sub>O<sub>2</sub> of 90% used for patients receiving supplemental oxygen at 15 liters/minute through non-rebreathing face mask; <sup>d</sup>Denominator of 201 patients receiving IMV; <sup>e</sup>Administration within 48 hours of initiation of invasive mechanical ventilation.

By April 14^th^, 2020, 33% of patients (86/257) had died following a median of 8 days in hospital (IQR 5-12) (Figure 1a). The median time to clinical deterioration following admission was 3 days (IQR 1-6). Only six deaths (7%; 6/86) occurred in patients under 50 years of age (Figure 1b). One-hundred-twenty-two (47%) remained hospitalized with a median duration of hospitalization 17 days (IQR 14-21). Forty-five (18%) were discharged alive, nine (20%) of which newly required supplemental oxygen, and 4 (2%) were transferred to another institution.

**Figure 1:**
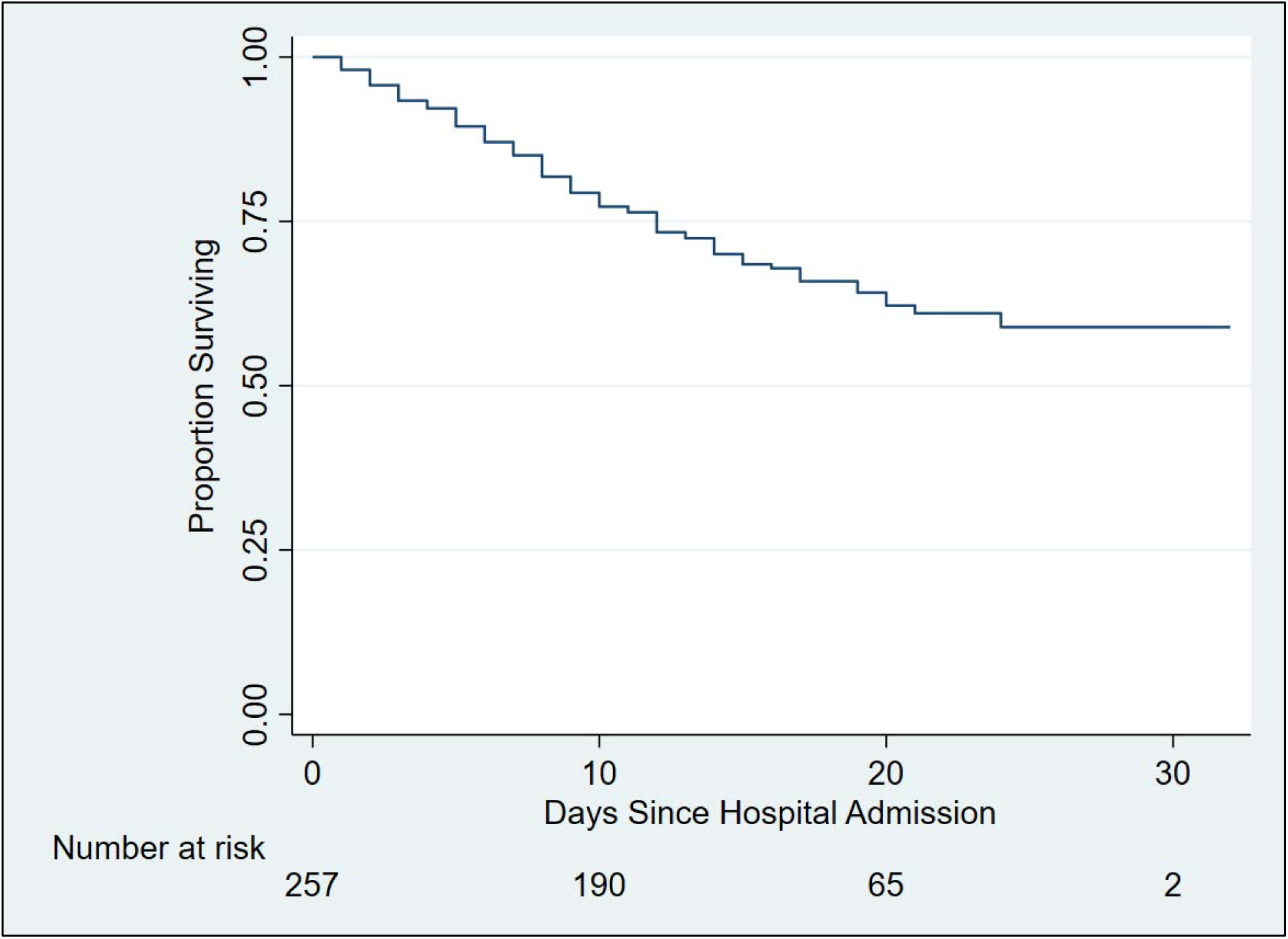
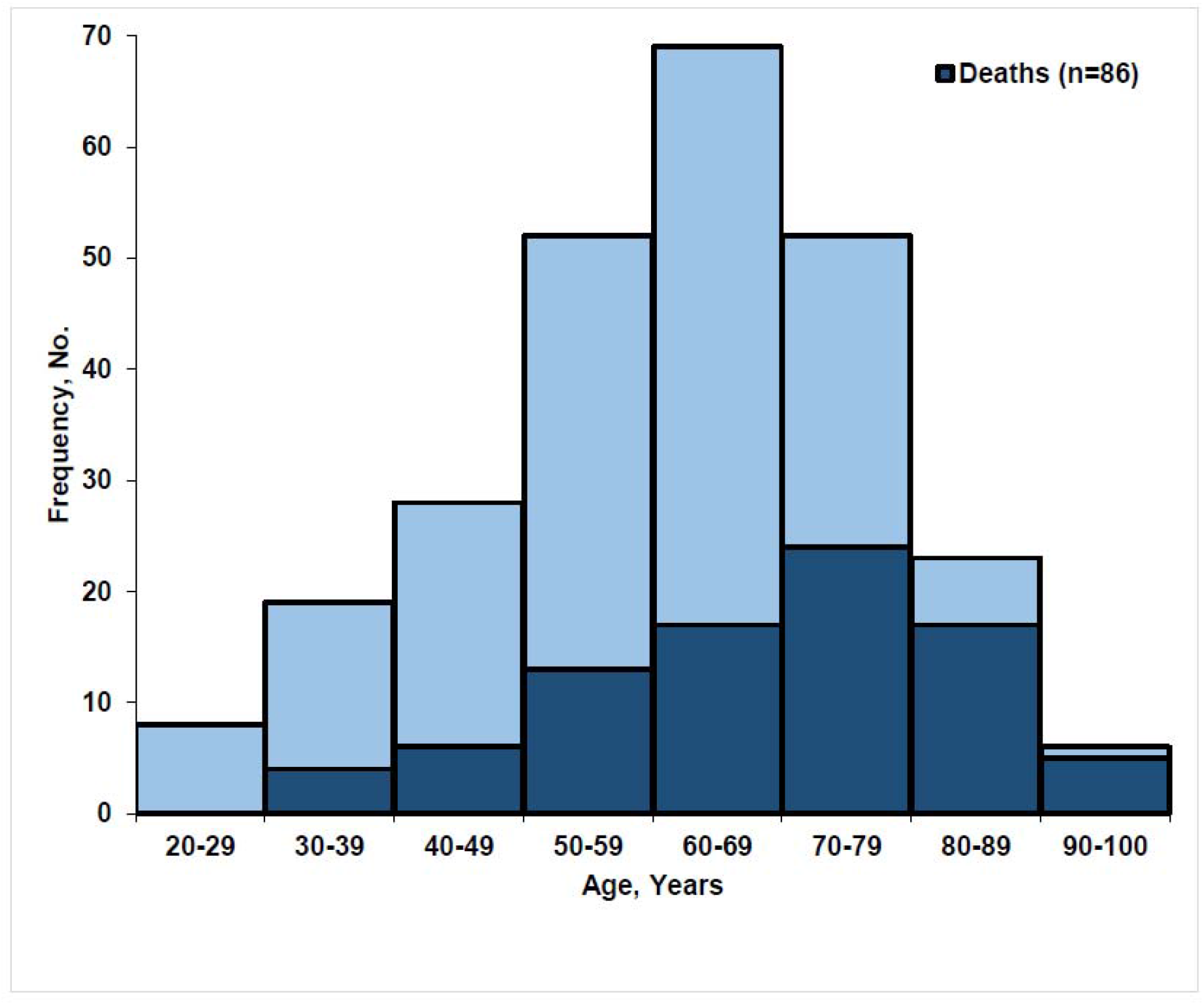
**(a)** Kaplan-Meier Survival Curve for Critically Ill Patients with COVID-19. **(b)** Age Distribution of Critically Ill Patients with COVID-19.

### Risk Factors for In-hospital Mortality

In multivariable Cox models (Table 4), older age (adjusted hazard ratio [aHR] 1.04, 95% confidence interval [CI] 1.02-1.06 per year), hypertension (aHR 2.12, 95% CI 1.13-3.99) (Figure S1 in data supplement), chronic obstructive pulmonary or interstitial lung disease (aHR 4.22, 95% CI 2.02-8.84), and higher concentrations of IL-6 (aHR 1.002, 95% CI 1.000-1.005 per one-unit increase) and d-dimer (aHR 1.010, 95% CI 1.003-1.018 per one-unit increase) at admission were associated with in-hospital mortality.

**Table 4:** Risk factors for in-hospital mortality.

| **Variable** | **Univariable HR**<br>**(95% CI)** | **Multivariable HR**<br>**(95% CI)** |
| --- | --- | --- |
| Age, per one-year increase | 1.05 (1.03-1.07) | 1.04 (1.02-1.06) |
| Male sex | 0.96 (0.61-1.50) | 1.35 (0.80-2.25) |
| Symptom duration prior to presentation, per day | 1.02 (0.98-1.07) | 1.09 (0.96-1.06) |
| Hypertension | 2.27 (1.37-3.79) | 2.13 (1.13-3.99) |
| Chronic cardiovascular disease | 0.62 (0.33-1.17) | 0.69 (0.34-1.42) |
| Chronic obstructive pulmonary and/or interstitial lung disease | 3.45 (2.00-5.97) | 4.22(2.02-8.84) |
| Chronic kidney disease | 1.58 (0.94-2.66) | 1.46 (0.81-2.13) |
| Diabetes mellitus | 0.89 (0.57-1.39) | 1.28 (0.77-2.13) |
| Body-mass-index $\geq 35$ | 0.74 (0.43-1.27) | 0.94 (0.55-1.77) |
| Interleukin-6, per one-unit increase | 1.001 (1.000-1.003) | 1.002 (1.000-1.005) |
| D-dimer, per one-unit increase | 1.011 (1.005-1.018) | 1.010 (1.003-1.018) |

Abbreviations: HR: hazard ratio

## Discussion

Among critically ill adults with COVID-19 admitted to two hospitals in New York City during the first 30-days of the city’s outbreak, the majority were men over 60 years with hypertension and diabetes mellitus, nearly half were obese, and 5% were HCWs. Over 75% of patients received IMV and high levels of PEEP, and nearly a third received RRT. As of April 14^th^, over 30% of patients had died in-hospital, and the rate of in-hospital death was associated with high-risk clinical factors as well as biomarkers of coagulopathy and dysregulated immune activation.

Over 20% of patients hospitalized with COVID-19 were critically ill with acute hypoxemic respiratory failure. This is consistent with reports from China,^3,4^ Italy,^5^ and preliminary U.S. data released by the Centers for Disease Control and Prevention,^16^ in which the incidence of ICU admission among patients admitted with COVID-19 ranged from 7-26%. This high incidence of critical illness among hospitalized patients has acute implications for U.S. hospital systems, specifically the potential need to increase ICU surge capacity in preparation for large numbers of patients requiring IMV and other forms of organ support.

Nearly 80% of patients received IMV during hospitalization for median durations of 5 and 14 days among non-survivors and survivors, respectively. This included 60% of patients who initially received less invasive methods of respiratory support. Although the proportion of patients in our cohort receiving IMV was higher than that reported in observational studies from China^3,4,6,7^ and Washington state,^9,10^ it is more similar to the rate recently reported from Italy,^8^ in which IMV was provided to 88% of critically COVID-19 patients. As in Italy, where the median ratio of P_a_O_2_ to F_i_O_2_ at ICU admission was 160,^8^ the higher proportion of patients requiring IMV in our cohort may be explained by the severity of hypoxemia, as the median nadir P_a_O_2_ to F_i_O_2_ in our population was 129. The sudden surge of critically ill patients admitted with severe ARDS initially outpaced our capacity to provide prone-positioning ventilation, which was only performed in three of eight ICUs at our institution at the start of the outbreak. We have since expanded our capacity for prone-positioning by deploying dedicated proning teams to all ICUs, including non-traditional ICU locations.

As of April 14^th^, 2020, over 30% of patients had died in the hospital. Similar to data reported elsewhere,^3,7,17^ we identified older age, hypertension, and chronic lung disease, as well as higher levels of d-dimer,^8^ as independent risk factors for poor outcomes. Higher levels of IL-6, which have been observed among COVID-19 patients with more severe clinical illness,^17,18^ were also associated with in-hospital mortality. Although the pathogenesis of severe COVID-19 remains incompletely understood, emerging data suggests that organ dysfunction and poor outcomes may be mediated by high levels of pro-inflammatory cytokines including IL-6 and dysregulated coagulation and thrombosis.^13,14,19^ Continued investigation of these pathological processes and the utility of their biomarkers is needed given recent reports of corticosteroid use and ongoing clinical trials of IL-6-receptor antagonists among COVID-19 inpatients, as well as rapidly evolving guidelines for anti-coagulation in this population.^17,20,21^

Consistent with unadjusted data from China,^3^ and Italy,^8^ hypertension was independently associated with poor in-hospital survival. Given the globally high burden of hypertension and emerging understanding of interactions between SARS-CoV-2 and angiotensin-converting-enzyme-2,^22^ further investigations are needed to better define a relationship, if any, between hypertension, exposure to renin angiotensin aldosterone system antagonists, and severe COVID-19.

Nearly 30% of patients in our cohort developed severe acute kidney injury requiring RRT during hospitalization. Consistent with emerging data from China,^23^ nearly 90% of evaluated patients had proteinuria detected at hospital admission. The high frequency of RRT in our patient population has considerable implications for resource allocation given limited supplies of RRT machines and consumables, as well as staffing requirements, necessary to provide continuous or intermittent RRT to critically ill patients. As the general incidence and underlying mechanisms of severe COVID-19-related kidney injury remain poorly understood,^23^ epidemiologic, clinical and biological investigations are necessary to inform hospital preparedness strategies and development of targeted preventive and treatment interventions.

Nearly half of the critically ill patients were obese. This observation is consistent with trends seen in critically ill COVID-19 patients from the United Kingdom,^24^ and in previous reports of adults with severe influenza,^25^ in which obesity has been associated with increased incidence of ICU admission and mortality. However, while obesity was more common in our adult patient population than the general New York City adult population (46% vs. 22%),^26^ we did not identify severe obesity (BMI ≥35) as an independent risk factor for mortality. Similar to other cardio-metabolic co-morbidities, further studies are needed to better define an interaction, if any, between obesity and either susceptibility to or severity of COVID-19.

Hydroxychloroquine or remdesivir, agents which have shown activity against SARS-CoV-2 *in vitro*,^27^ were administered to over 70% of patients. Although the safety and efficacy of remdesivir among patients with COVID-19 remains uncertain, a recent uncontrolled observational study reported clinical improvement in 68% (36/53) of patients with severe COVID-19 who were administered the agent through compassionate use access.^28^ However, adverse events were reported in 60% of patients, most commonly elevated transaminases. As the safety and efficacy of hydroxychloroquine in severe COVID-19 are unknown, the National Institutes of Health recently launched a blinded, placebo-controlled clinical trial among hospitalized patients with COVID-19 in the U.S.^29^ In the absence of published clinical trial data for hydroxychloroquine and remdesivir, use of these agents should be decided on a case-by-case basis.

Five percent of critically ill patients were HCWs. Although nosocomial SARS-CoV-2 infection cannot be determined with certainty given widespread community transmission, COVID-19-related critical illness in these individuals highlights the risks facing frontline HCWs, such as in Italy, where over 15,000 HCWs have been infected as of April 11^th^, 2020.^30^ Continued and consistent access to personal protective equipment for hospital staff is imperative to prevent nosocomial transmission, optimize HCW safety, and ensure an adequate workforce.

This study has many strengths. First, our study represents the largest cohort of patients with COVID-19-related critical illness reported to-date in the United States. Second, as patients were identified and data were collected prospectively, our findings reflect the ongoing outbreak of COVID-19 in New York City, currently the largest in the world. Third, we collected data using a globally harmonized, World Health Organization-endorsed case record form. Fourth, we augmented collection of standard clinical and laboratory data with clinically- and pathologically-relevant biomarkers, concentrations of which were available for nearly all patients. Lastly, given the prospective nature of our study, our analyses were performed with near-complete data, with in-hospital outcomes known for all included patients through April 14^th^, 2020.

This study also has several limitations. First, our study was conducted in two academic hospitals in northern Manhattan, potentially limiting generalizability to hospital settings elsewhere in New York City, especially in terms of the demographic characteristics of the patient population. However, our sites included both a large tertiary referral hospital and a smaller, community-based hospital, thereby increasing generalizability to other clinical settings. Second, our analyses incorporated data collected through April 14^th^, 2020. As definitive in-hospital vital status is not yet known for patients who remained hospitalized after this date, the 33% of inpatient deaths reported here represent the minimum in-hospital case-fatality-rate for our cohort. Third, patients presented to the hospital at varying times in their illness course, which may have impacted their clinical course and outcomes. To mitigate the potential impact of this variance on our analyses, we included time from symptom onset to presentation as a co-variable in our regression models. Fourth, of available biomarkers, we included IL-6 and d-dimer in our multivariable models given pathophysiologic and treatment implications. We did not analyze serial concentrations of these and other biomarkers, which may fluctuate considerably over the course of illness.

In conclusion, critical illness among patients hospitalized with COVID-19 in New York City is common and associated with a high frequency of invasive mechanical ventilation, extra-pulmonary organ dysfunction, and substantial in-hospital mortality.

## Data Availability

Data are available from the corresponding author upon reasonable request.

## Acknowledgements

The authors would like to thank their patients and fellow healthcare workers for providing outstanding patient care at considerable personal risk.

## Data Supplement

**Table S1:** Ordinal Scale Used to Evaluate Clinical Status During Hospitalization.

| **Clinical Status** | **Point** |
| --- | --- |
| Not hospitalized, performing normal activities | 1 |
| Not hospitalized, but unable to perform normal activities | 2 |
| Hospitalized, not requiring supplemental oxygen | 3 |
| Hospitalized, requiring supplemental oxygen via nasal cannula | 4 |
| Hospitalized, requiring respiratory support via non-rebreather mask, or high-flow nasal cannula oxygen therapy, or noninvasive mechanical ventilation | 5 |
| Hospitalized, requiring extracorporeal membrane oxygen, invasive mechanical ventilation, or both | 6 |
| Death | 7 |

**Figure S1:**
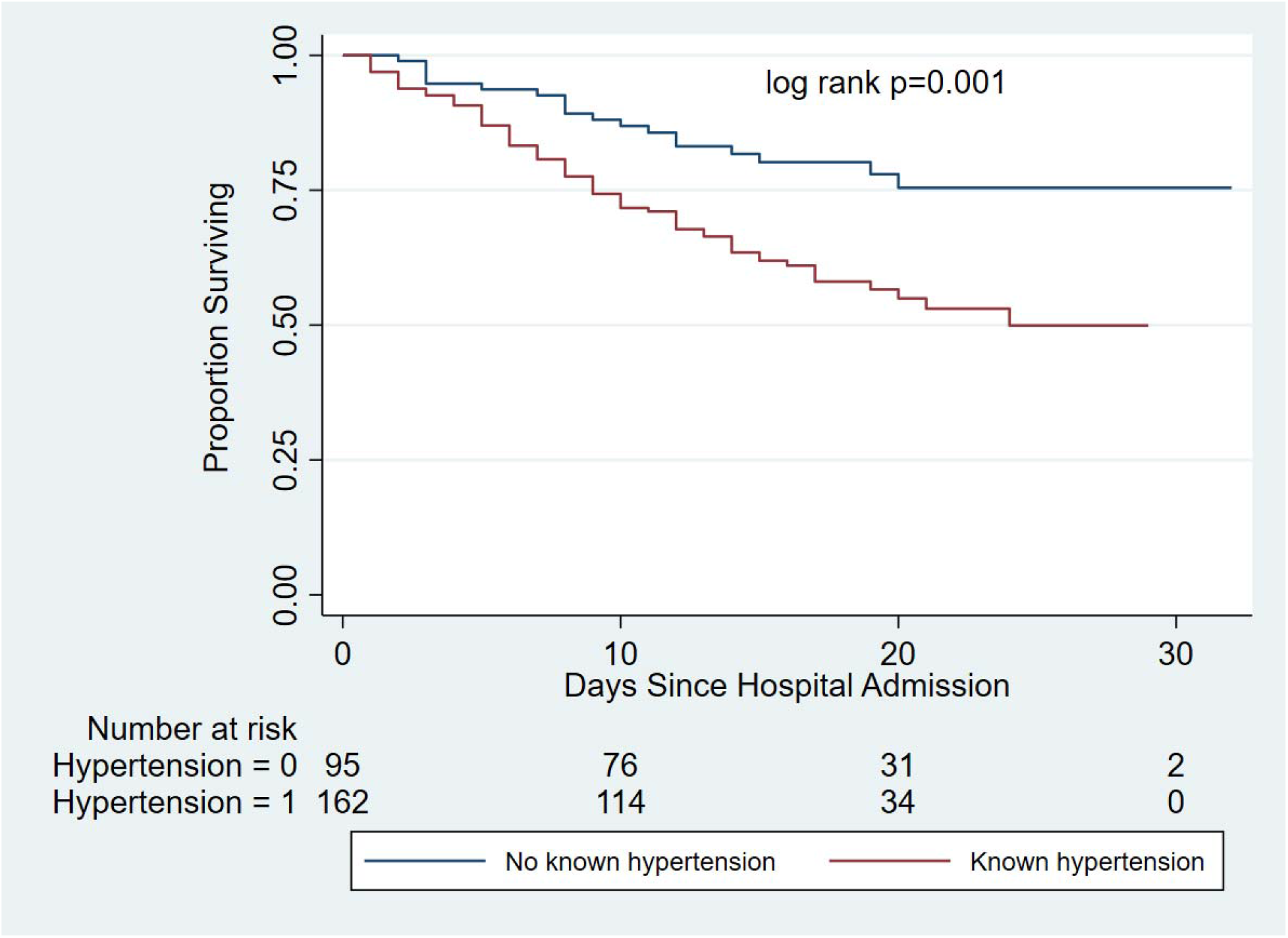
Unadjusted Kaplan-Meier Survival Curves by Hypertension Diagnosis Among Critically Ill Patients with COVID-19.

## References

1) Coronavirus disease-2019: cases in U.S. Atlanta: U.S. Centers for Disease Control and Prevention, April 11, 2020. (https://www.cdc.gov/coronavirus/2019-ncov/cases-updates/cases-in-us.html). Accessed April 11th, 2020.

2) COVID-19 Tracker. Albany: New York State Department of Health, April 11, 2020. (https://covid19tracker.health.ny.gov/views/NYS-COVID19-Tracker/NYSDOHCOVID-19Tracker-Map?%3Aembed=yes&%3Atoolbar=no&%3Atabs=n). Accessed April 11th, 2020.

3) Wu Z, McGoogan JM. Characteristics of and Important Lessons From the Coronavirus Disease 2019 (COVID-19) Outbreak in China: Summary of a Report of 72D314 Cases From the Chinese Center for Disease Control and Prevention. JAMA 2020; doi: 10.1001/jama.2020.2648.

4) Guan WJ, Ni ZY, Hu Y, et al. Clinical Characteristics of Coronavirus Disease 2019 in China. N Engl J Med 2020; doi: 10.1056/NEJMoa2002032.

5) Grasselli G, Pesenti A, Cecconi M. Critical Care Utilization for the COVID-19 Outbreak in Lombardy, Italy: Early Experience and Forecast During an Emergency Response. JAMA 2020; doi: 10.1001/jama.2020.4031.

6) Yang X, Yu Y, Xu J, et al. Clinical course and outcomes of critically ill patients with SARS-CoV-2 pneumonia in Wuhan, China: a single-centered, retrospective, observational study. Lancet Respir Med 2020; doi: 10.1016/S2213-2600(20)30079-5.

7) Zhou F, Yu T, Du R, et al. Clinical course and risk factors for mortality of adult inpatients with COVID-19 in Wuhan, China: a retrospective cohort study. Lancet 2020; 395:1054–1062.

8) Grasselli G, Zangrillo A, Zanella A, et al. Baseline Characteristics and Outcomes of 1591 Patients Infected With SARS-CoV-2 Admitted to ICUs of the Lombardy Region, Italy. JAMA 2020; doi: 10.1001/jama.2020.5394.

9) Arentz M, Yim E, Klaff L, et al. Characteristics and Outcomes of 21 Critically Ill Patients With COVID-19 in Washington State. JAMA 2020; doi: 10.1001/jama.2020.4326.

10) Bhatraju PK, Ghassemieh BJ, Nichols M, et al. Covid-19 in Critically Ill Patients in the Seattle Region - Case Series. N Engl J Med 2020; doi: 10.1056/NEJMoa2004500.

11) ISARIC-WHO COVID-19 Case Record Form. Oxford: International Severe Acute Respiratory and Emerging Infection Consortium. https://isaric.tghn.org/COVID-19-CRF/. Accessed March 2nd, 2020.

12) Research and development blueprint: novel coronavirus. Geneva: World Health Organization, 2020. https://www.who.int/blueprint/priority-diseases/key-action/COVID-19_Treatment_Trial_Design_Master_Protocol_synopsis_Final_18022020.pdf. Accessed April 11th, 2020.

13) Mehta P, McAuley DF, Brown M, Sanchez E, Tattersall RS, Manson JJ. COVID-19: consider cytokine storm syndromes and immunosuppression. Lancet 2020; 395:1033–1034.

14) Tang N, Bai H, Chen X, Gong J, Li D, Sun Z. Anticoagulant treatment is associated with decreased mortality in severe coronavirus disease 2019 patients with coagulopathy. J Thromb Haemost 2020; doi.org/10.1111/jth.14817.

15) Kelly CA, Upex A, Bateman DN. Comparison of consciousness level assessment in the poisoned patient using the alert/verbal/painful/unresponsive scale and the Glasgow Coma Scale. Ann Emerg Med. 2004;44:108–13.

16) U.S. CDC COVID-19 Response Team. Severe Outcomes Among Patients with Coronavirus Disease 2019 (COVID-19) - United States, February 12-March 16, 2020. MMWR Morb Mortal Wkly Rep 2020; 69:343–346.

17) Wu C, Chen X, Cai Y, et al. Risk Factors Associated With Acute Respiratory Distress Syndrome and Death in Patients With Coronavirus Disease 2019 Pneumonia in Wuhan, China. JAMA Intern Med 2020; doi: 10.1001/jamainternmed.2020.0994.

18) Ruan Q, Yang K, Wang, W, et al. Clinical predictors of mortality due to COVID-19 based on an analysis of data of 150 patients from Wuhan, China. Intensive Care Med 2020; doi.org/10.1007/s00134-020-05991-x.

19) Zhou Y, Fu B, Zheng X, et al. Pathogenic T cells and inflammatory monocytes incite inflammatory storm in severe COVID-19 patients. Natl Sci Rev 2020; doi:10.1093/nsr/nwaa041.

20) Evaluation of the Efficacy and Safety of Sarilumab in Hospitalized Patients with COVID-19. Bethesda: U.S. National Library of Medicine. Clinicaltrials.gov Identifier NCT04315298. (https://clinicaltrials.gov/ct2/show/NCT04315298). Accessed April 11th, 2020.

21) Thachil J, Tang N, Gando S, et al. International Society on Thrombosis and Haemostasis interim guidance on recognition and management of coagulopathy in COVID-19. J Thromb Haemost 2020; https://doi.org/10.1111/jth.14810.

22) Zheng, Y., Ma, Y., Zhang, J. et al. COVID-19 and the cardiovascular system. Nat Rev Cardiol 2020; doi.org/10.1038/s41569-020-0360-5

23) Naicker S, Yang CW, Hwang SJ, Liu BC, Chen JH, Jha V. The Novel Coronavirus 2019 epidemic and kidneys. Kidney Int 2020; doi: 10.1016/j.kint.2020.03.001.

24) ICNARC report on COVID-19 in critical care. London: Intensive Care National Audit & Research Centre, April 10th, 2020. (https://www.icnarc.org/Our-Audit/Audits/Cmp/Reports). Accessed April 11th, 2020.

25) Fezeu L, Julia C, Henegar A, et al. Obesity is associated with higher risk of intensive care unit admission and death in influenza A (H1N1) patients: a systematic review and meta-analysis. Obes Rev 2011;12:653–9.

26) New York State Community Health Indicator Reports. Albany: New York state Department of Health, April 1, 2019. (https://webbi1.health.ny.gov/SASStoredProcess/guest?_program=%2FEBI%2FPHIG%2Fapps%2Fchir_dashboard%2Fchir_dashboard&p=sh&stop=10). Accessed April 11th, 2020.

27) Wang M, Cao R, Zhang L. et al. Remdesivir and chloroquine effectively inhibit the recently emerged novel coronavirus (2019-nCoV) in vitro. Cell Res 2020; 30: 269–271

28) Grein J, Ohmagari N, Shin D, et al. Compassionate Use of Remdesivir for Patients with Severe Covid-19. N Engl J Med. 2020; doi: 10.1056/NEJMoa2007016.

29) Outcomes Related to COVID-19 Treated With Hydroxychloroquine Among In-patients With Symptomatic Disease (ORCHID). Bethesda: U.S. National Library of Medicine. ClinicalTrials.gov Identifier: NCT04332991. (https://clinicaltrials.gov/ct2/show/NCT04332991).

30) Integrated surveillance of COVID-19 in Italy. Rome: EpiCentro. (https://www.epicentro.iss.it/en/coronavirus/bollettino/Infografica_10aprile%20ENG.pdf). Accessed April 11th, 2020.

